# Development and validation of an automated radiomic CT signature for detecting COVID-19

**DOI:** 10.1101/2020.04.28.20082966

**Authors:** J. Guiot, A. Vaidyanathan, L. Deprez, F. Zerka, L. Danthine, A.N. Frix, M. Thys, M. Henket, G. Canivet, S. Mathieu, E. Eftaxia, P. Lambin, N. Tsoutzidis, B. Miraglio, S. Walsh, M. Moutschen, R. Louis, P. Meunier, W. Vos, R.T.H. Leijenaar, P. Lovinfosse

**Author notes:** Guiot J. & Vaidyanahan A. have equally contributed as first author. Lovinfosse P. & Leijenaar R.T.H. have equally contributed as last author. **Corresponding Author:** Dr. Julien Guiot, Pneumology department, C.H.U. - SART TILMAN - B35, 4000 Liège, BELGIUM.

## Abstract

**Background:** The coronavirus disease 2019 (COVID-19) outbreak has reached pandemic status. Drastic measures of social distancing are enforced in society and healthcare systems are being pushed to and over their limits.

**Objectives:** To develop a fully automatic framework to detect COVID-19 by applying AI to chest CT and evaluate validation performance.

**Methods:** In this retrospective multi-site study, a fully automated AI framework was developed to extract radiomics features from volumetric chest CT exams to learn the detection pattern of COVID-19 patients. We analysed the data from 181 RT-PCR confirmed COVID-19 patients as well as 1200 other non-COVID-19 control patients to build and assess the performance of the model. The datasets were collected from 2 different hospital sites of the CHU Liège, Belgium. Diagnostic performance was assessed by the area under the receiver operating characteristic curve (AUC), sensitivity and specificity.

**Results:** 1381 patients were included in this study. The average age was 64.4±15.8 and 63.8±14.4 years with a gender balance of 56% and 52% male in the COVID-19 and control group, respectively. The final curated dataset used for model construction and validation consisted of chest CT scans of 892 patients. The model sensitivity and specificity for detecting COVID-19 in the test set (training 80% and test 20% of patients) were 78.94% and 91.09%, respectively, with an AUC of 0.9398 (95% CI: 0.875–1). The negative predictive value of the algorithm was found to be larger than 97%.

**Conclusions:** Benchmarked against RT-PCR confirmed cases of COVID-19, our AI framework can accurately differentiate COVID-19 from routine clinical conditions in a fully automated fashion. Thus, providing rapid accurate diagnosis in patients suspected of COVID-19 infection, facilitating the timely implementation of isolation procedures and early intervention.

## Introduction

The rapid outbreak of coronavirus disease 2019 (COVID-19), originating from severe acute respiratory syndrome coronavirus 2 (SARS-COV-2) infection, has plainly become a public health emergency of international concern [1]. COVID-19 has created worldwide an immense adverse impact. Globally there has been 1,131,713 confirmed cases and 60,115 deaths as of April 4^th^ 2020 [2].

The disease is currently confirmed by reverse-transcription polymerase chain reaction (RT-PCR) [3]. There is, however, evidence that the sensitivity of RT-PCR may not be optimal for the objective of very early detection and early intervention of COVID-19 patients [4]. Due to the limited supply of RT-PCR kit, the lengthy turnaround times, and the emergence of false-negative nucleic acid cases, some experts propose to diagnose suspected cases using the widely available time-saving and non-invasive imaging approach of chest computed tomography (CT) rather than RT-PCR [5,6] as CT can capture imaging features from the lung associated with COVID-19 [7] early in the course of the disease [8]. CT could thus serve as an efficient and effective way to screen, diagnosis, and possibly triage COVID-19 patients. Despite these apparent strengths, there are notable calls of caution with respect to using CT for these purposes [9,10], due to increased radiation exposure of the population and the risk of cross-infection if disinfection is not properly implemented.

Nevertheless, if chest CT is to be utilized in this pandemic it needs to be applied optimally. Artificial intelligence (AI) using machine learning technology in the medical imaging domain has accomplished impressive results due to the intrinsic properties of machine vision [11–14] and can be leveraged in this crisis.

Radiomics is the high-throughput mining of quantitative image features from standard-of care medical imaging that enables data to be extracted and applied within clinical decision support systems to improve diagnostic, prognostic, and/or predictive accuracy [15]. Conceptually, radiomics is a bridge between imaging and precision medicine [16].

In this study, we hypothesize that a radiomics analysis can identify a diagnostic signature for COVID-19 infection, based on standard of care chest CT imaging. As a result, we present a fully automated AI framework to detect COVID-19 using chest CT, referred to as COVIA (‘coronavirus intelligence artificielle’). Other non-COVID-19 patients were included as controls to test the robustness of the model performance.

## Materials and methods

### Ethics

The study has been approved by the local ethics committee of the CHU-Liège (EC number 116/2020). The institutional review board waived the requirement to obtain written informed consent for this retrospective case series, since all analyses were performed on de-identified (i.e., anonymized) data and there was no potential risk to patients.

### Subjects

Two cohorts of patients were included retrospectively in this study. Cohorts came from two sites (CHU Sart-Tilman and CHU Notre Dame des Bruyères) in Liège, Belgium. The first cohort (label: COVID) consists of all patients with COVID-19 infection confirmed by RT-PCR that underwent chest CT imaging before March 28^th^, 2020. The second cohort (label: Control) consisted of consecutive patients that underwent chest CT imaging between October 1^st^, 2019 and October 24^th^, 2019, which ensures that none of these patients were infected by COVID-19. No other inclusion or exclusion criteria were considered while collecting the data. This resulted in a set of CT images from either COVID-19 infected patients or non-infected patients (normal and a variety of diseases) representing real life conditions.

### Radiomics

Radiomics is an in-depth automatic analysis that aims to quantify all the information contained in medical images. Some of this information might go undetected for by the naked eye but is present in the image and, when detected by radiomics, opens a pathway towards precision imaging medicine. The schematic representation in Figure 1 depicts the radiomics workflow applied in this study. The following sections will detail each step in the workflow.

**Figure 1.**
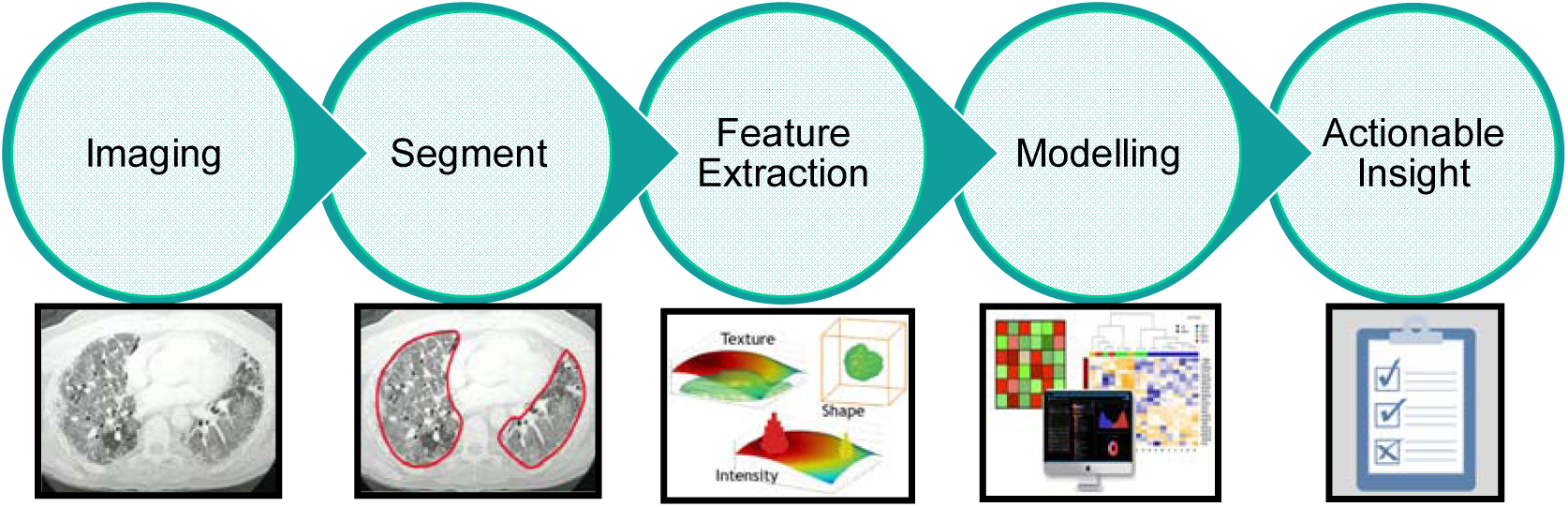
The radiomics workflow applied in this study: Schematic representation of the steps performed to produce a signature to support clinical decision making.

### Imaging

All CT images used in the study were acquired on one of five multidetector CT scanners (Siemens Edge Plus (2), GE Revolution CT (1), GE Brightspeed (2)) available at the sites. Since CT images were collected retrospectively, no standardized scan protocol was available over the complete dataset. In order to prevent excess variability in the imaging used for model generation the following criteria for radiomic analysis was used:

- Lungs completely visible in the scan
- Slice increment less than 1.5 mm
- No missing slices
- For GE scans: STANDARD reconstruction kernel
- For Siemens scans: B30-range reconstruction intervals

### Lung segmentation

The lungs were segmented as a single structure using RadiomiX (OncoRadiomics SA, Liège, Belgium) based on convolutional neural networks by combining 2D and 3D architectures. The predicted segmentations of each architecture are ensembled and the intersection constitutes the final lung segmentation which is used for extraction of radiomics features. Figure 2 shows example segmentations for two four patients from both the COVID and Control groups. Complete details on the segmentation methods can be found in the online supplement.

**Figure 2.**
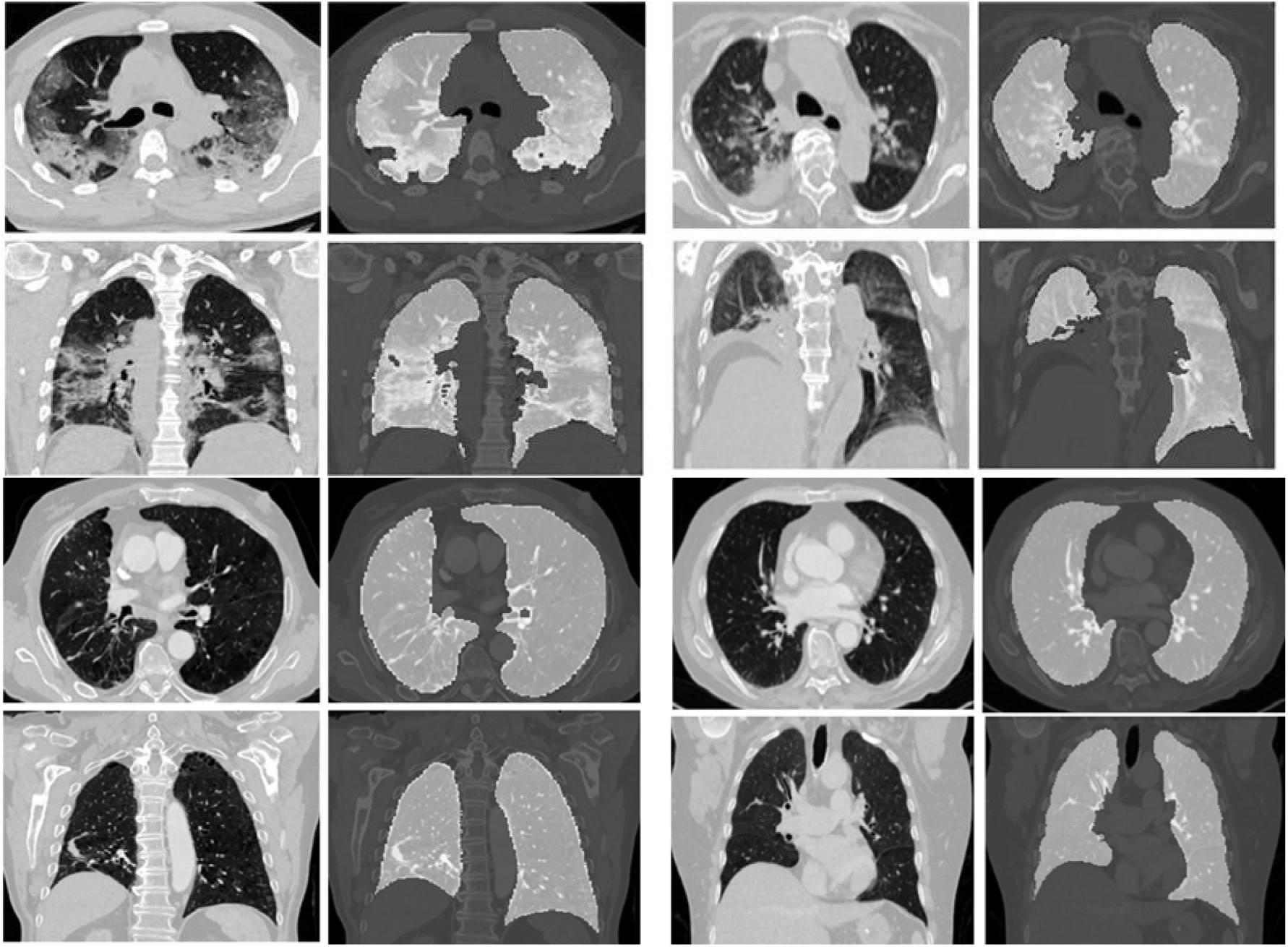
Axial and coronal slices with accompanying segmentation masks. Top left quadrant - Typical aspect of COVID-19 pneumonia characterized by bilateral multilobe ground-glass opacities of peripheral/subpleural distribution, with intralesional reticulations, presenting of a “crazy paving” aspect. Also found are subpleural condensations and retraction bronchiectasis, typical of organizing pneumonia; Top right quadrant - Atypical aspect of COVID-19 pneumonia, with posterior right lower lobe condensation and retraction of the ipsilateral diaphragm. Central and peripherical ground-glass opacities in right lower lobe, right upper lobe and left upper lobe; Bottom left quadrant - Typical chronic obstructive pulmonary disease COPD chest CT characterized by severe centrilobular and para-septal emphysema, associated with cylindrical bronchiectasis and bronchial walls thickening. Right peripherical upper lobe tree in bud pattern seen in bronchiolitis. Middle lobe crescent-shaped atelectasis condensation; Bottom right quadrant - Normal chest CT.

### Feature Extraction

For each patient, 166 image features were extracted from the lung segmentation using RadiomiX (OncoRadiomics SA, Liège, Belgium) based upon quantitative image analysis technology. The extracted features comprised first order statistics, texture, and shape. A bin width of 25 Hounsfield unit was used for image intensity discretization. Mathematical descriptions of all features are described in [16].

### Modelling

A statistical model that can provide a binary classification of patients (COVID-19 infected or not) has been trained and validated using a regularized logistic regression model [17].

Stratified partitioning was performed to randomly split the data into 80% for model training and 20% for validation. Stratification was performed for age, gender, diagnosis of COPD and COVID-19 status, ensuring a balanced distribution of these characteristics in both data partitions.

Multivariable logistic regression with Elastic Net regularization was performed in the training data set. Highly correlated features, features with near zero variance and linear combinations between features were first eliminated from further analysis. For each highly correlated feature pair (Pearson correlation coefficient ρ > 0.9), the variable with the largest mean absolute correlation with all remaining features was removed. Model training was performed using 100 times repeated 10-fold cross-validation to select the optimal model hyperparameters, optimizing for the area under the receiver operator curve (AUC). All features were standardized before modelling. Class weighting was applied to address for class imbalance in the training data. To further reduce the chance of overfitting to the training data, we selected the simplest candidate model (i.e., the model with fewest non-zero coefficients) within one standard error of the best performing model.

Model performance was validated in the test data partition. Here, the AUC was used to assess model performance in discriminating between COVID-19 positive and COVID-19 negative patients. Classification performance was assessed with sensitivity and specificity analysis. All statistical analysis was performed in R (version 3.6.2).

## Results

### Study population

Table 1 lists the study population characteristics for the COVID and Control cohorts, as well as the main CT findings as scored by radiologists. Both cohorts have a similar mean age and male/female distribution. For the COVID-19 infected patients 69% needed O2 at admission, resulting in 37% of patients ending up in the ICU. 17% of COVID-19 patients needed mechanical ventilation and 4% died.

**Table 1:** Summary of patient characteristics (age, gender and CT findings scored by radiologist) per cohort.

|  | CONTROL (n=1200) | COVID (n=181) |
| --- | --- | --- |
| Age (years) | 63.8 ± 14.4 | 64.4±15.8 |
| Gender (% Male) | 52% | 56% |
| Normal (%) | 33% | 4.41% |
| Neoplasia (%) | 8.73% | 0% |
| CAP (%) | 12.50% | 8.10% |
| COPD (%) | 26% | 19.33% |
| Isolated pleurisy (%) | 6.2% | 1.10% |
| Pulmonary embolism (%) | 0.77% | 1.10% |
| Nodule (%) | 19% | 6.62% |
| Chronic inflammation* (%) | 8.48% | 5.52% |
| Pneumothorax (%) | 0.68% | 0% |
| Isolated atelectasis (%) | 3.68% | 3.31% |

### Data curation

After an automated quality check on the inclusion criteria, CT images and lung segmentations for a total number of 892 patients were included for further processing. A flow chart describing the overall workflow from data collection to model training and testing is shown in Figure 3.

**Figure 3.**
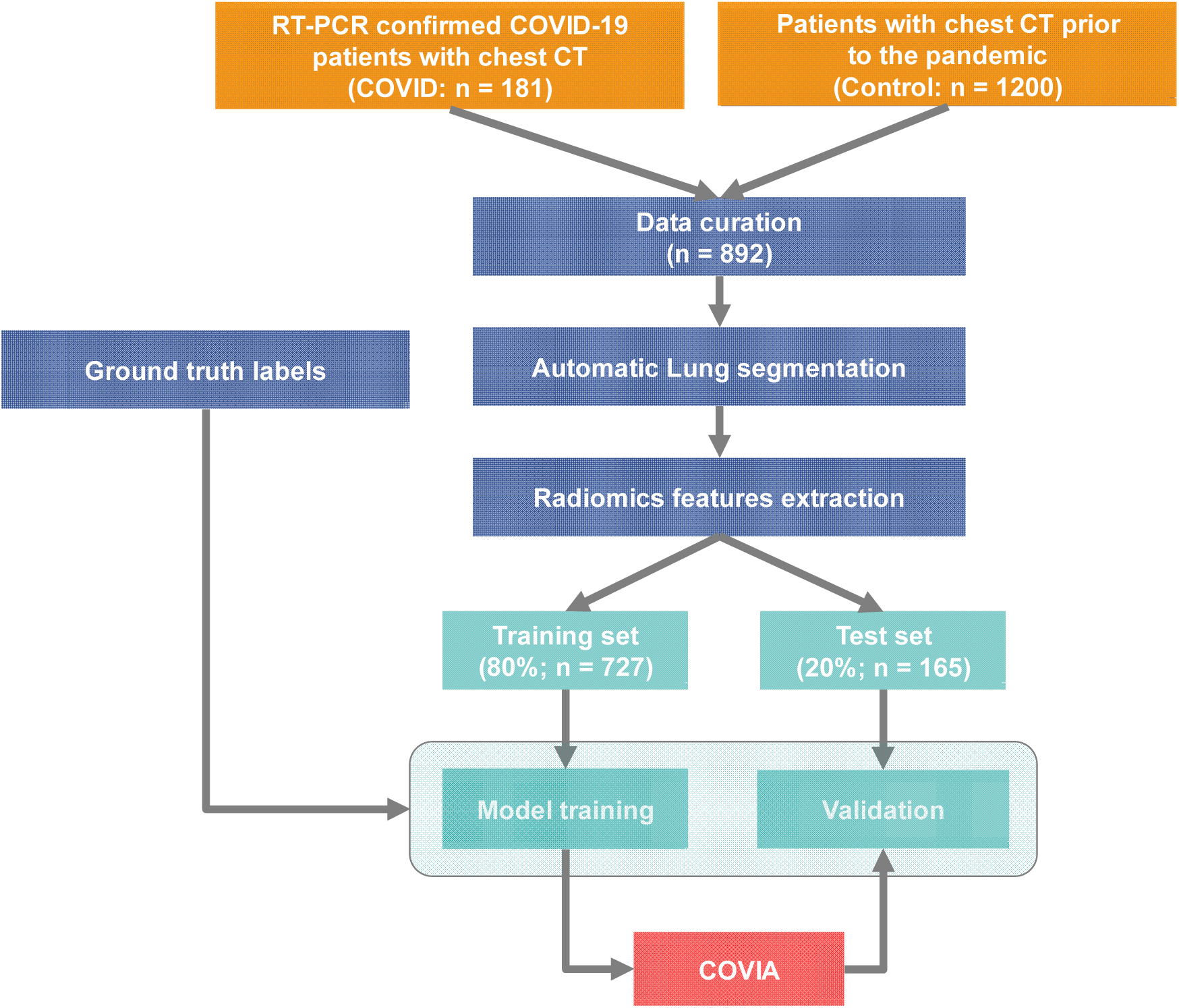
Flow diagram: Data was collected and combined from the COVID and Control cohorts. Lungs were segmented from the combined dataset and radiomics features were extracted. The Radiomics data was split into a training set for COVIA model development and a test set to validate the performance of COVIA with unseen patient CTs.

### COVID-19 infection status prediction

The final prediction model included 30 radiomics features with a non-zero regression coefficient. In the test data partition, the sensitivity and specificity for COVID-19 classification are 78.9% and 91.1%, respectively. The overall accuracy is 89.7% (95% CI: 84 – 93.9). The positive predictive value and negative predictive value are 53.6% and 97.1%, respectively. The ROC curve is shown in Figure 4. The corresponding AUC value for discriminating between COVID-19 positive and negative cases is 0.939 (95% CI: 0.875–1).

**Figure 4.**
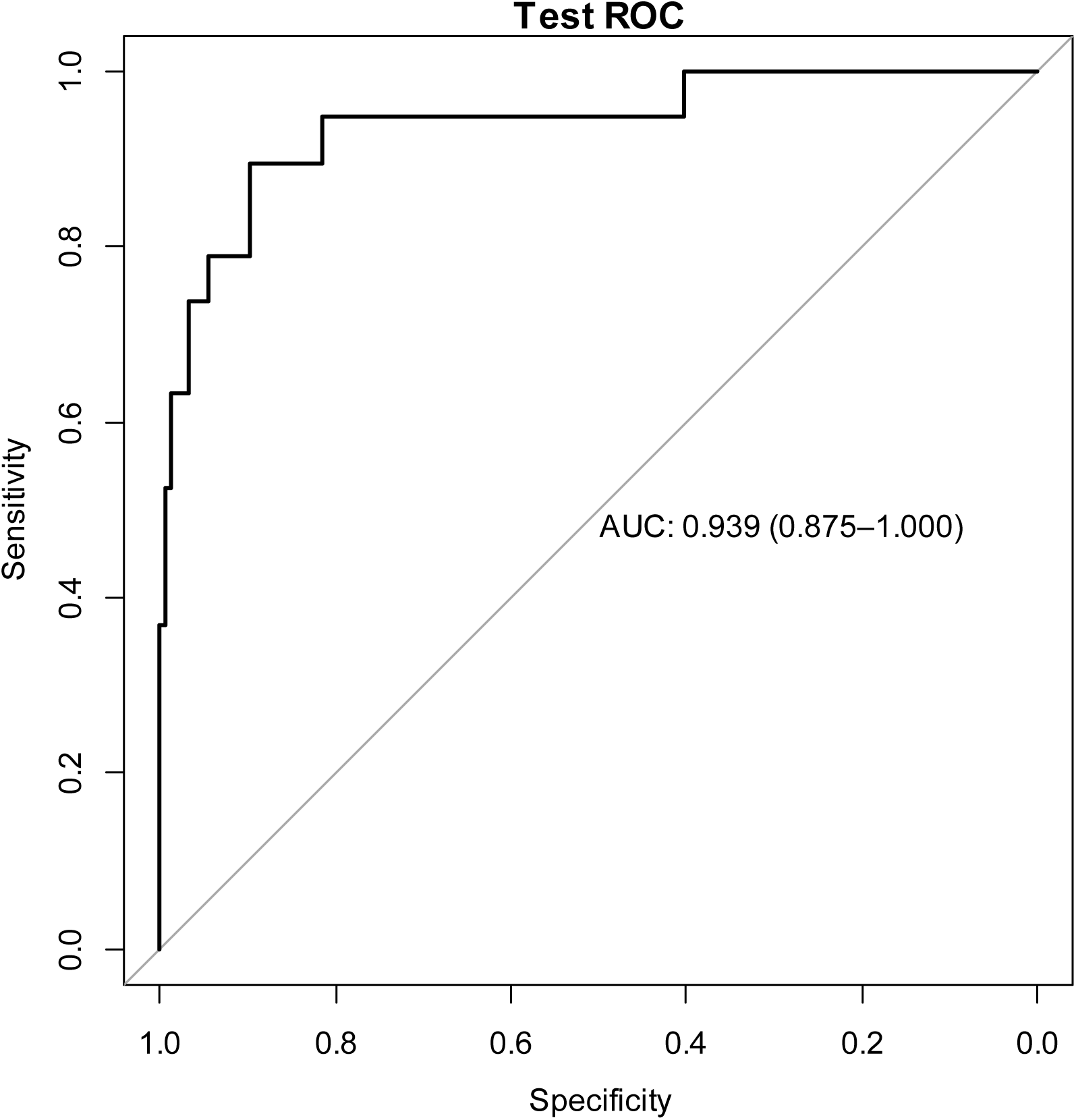
ROC curve. The ROC plot illustrates the performance (black curve) of the AI framework to discriminate between COVID-19 positive and negative cases in the test set (20% data partition) with an AUC of 0.939 (95% CI: 0.875–1).

## Discussion

COVID-19 has spread rapidly across the globe and rate of infection is accelerating. Therefore, rapid early diagnosis of the disease is essential for early intervention and swift isolation of patients in order to prevent the spread of the virus. RT-PCR is considered as the ‘gold standard’, however, there are reports of false-negatives occurring which are eventually confirmed as true-positive by repeated swab tests [18]. False negatives can be a significant problem in high-throughput settings operating under severe pressure [19]. The correct operation of the test is crucial and there is ambiguity with respect to the kinetics of SARS-CoV-2 viral shedding, thus the timing of the test may very well dictate the result. Furthermore, it is also unclear what kind of clinical sample is most appropriate as nasopharyngeal swabs may offer greater consistency than sputum samples. When considering the limited supply of RT-PCR, the growing backlog and the likely increasing pressure and turnaround times in laboratories, the issues pertaining to false-negatives, some experts have concluded that to diagnose suspected cases using the widely available time-saving and non-invasive imaging approach of chest CT is justified as this approach can sensitively and specifically identify COVID-19 patients. The high NPV (>97%) of our model provides further justification of using CT imaging-based diagnosis a primary tool for COVID-19 patient management.

Whereas similar studies in COVID-19 focus mainly on the detection of various diseased regions (including ground-glass opacification, consolidation, bilateral involvement, peripheral and diffuse distribution amongst others) in the lung [20,21] [22], our approach performs an easy segmentation of the lungs as one single structure, which is by far an easier task to automate with AI. Features for our quantitative image analysis are extracted from this whole lung structure and subsequently used for prediction model application and COVID-19 infection status classification. In the end this constitutes a fully automated clinical decision support tool for the diagnosis of COVID-19, which is able to provide an objective, robust (i.e., no user variability) and easy to interpret classification (yes-no) of COVID-19 infection status. The complete workflow takes between 40–60s and is thus, providing a rapid and accurate diagnosis in patients with suspected of COVID-19 infection, facilitating the timely implementation of isolation procedures and early intervention. For 160 CT scans of the patients in the COVID group a radiologist interpretation of COVID status was also available. Out of these patients, 123 were diagnosed as COVID-19 infected patients based on the image analysis from the radiologists (sensitivity=0.77). It is important to mention that our AI algorithm has similar sensitivity as compared to a trained radiologist, but our results were obtained in a large dataset with a COVID-19 prevalence of only 11% while the radiologist scoring happened in the midst of the current pandemic where COVID-19 prevalence was greatly elevated amongst the patients undergoing a chest CT exam. We therefor expect that our algorithm will outperform radiologists once the pandemic phase of COVID-19 is over. This hypothesis will be tested in a virtual clinical trial setting in the coming weeks. This aspect is vital in the context of incidental findings, which is of increasing relevance [23]. An automated AI solution is helpful in assisting the accurate identification of potentially COVID-19 positive patients, alerting the radiology department that a ‘clean machine’ now requires decontamination.

One argument against this image-based screening approach is the increased radiation exposure of the population. It is certainly important to keep the radiation dose to the public arising from medical examinations as low as reasonably achievable [24]. However, when we consider the exemplars of breast and lung cancer (two indications with established or soon to be established ionizing radiation medical imaging screening programs [25,26]) the expected mortality rate linked to the disease in the US this year is approximately 42,000 and 135,000 respectively [27,28]. A recent estimate of the mortality rate in the US this year due to COVID-19 was between 1.1-2.2 million [29]. These estimates would seem to support the justification to use CT as a screening tool for COVID-19. Moreover, a recent study reported that no effect on human DNA was detected when low dose CT procedures are used [30].

Another objection is the risk of cross-infection if disinfection of imaging equipment is not properly implemented. This is of great important because one of the great benefits of early diagnosis is the ability to swiftly isolate the patient and thus limit the spread of the virus. Thus, if the CT scanner were to become a vector for infection this would completely negate any benefit accrued. It is therefore vital that radiology departments are adequately prepared for managing (suspected) COVID-19 patients in a safe, efficient, and effective fashion [31].

A general objection of AI methods is the lack of transparency and interpretability. This is not the case with our approach, as ‘handcrafted’ radiomics features are explicitly defined and linked to clearly specified regions of interest within the images, driving the decision of the algorithm. Thus, clearly and intelligibly quantifying the image phenotype, which has also been shown to provide a means of connecting to the underlying biology [32].

We have encountered some limitations in this study. Firstly, COVID-19 is caused by a SARS-CoV-2 and may have similar imaging characteristics as pneumonia caused by other types of viruses. However, due to the lack of laboratory confirmation of the aetiology for each of these cases, we were not able to select other viral pneumonias for comparison in this study. Although our Control group of non-COVID-19 patients contains several patients (CAP in Table 1, 12.5%) with pneumonia (either viral, bacterial or organizing pneumonia from any cause), it would be desirable to test the performance of our algorithm in distinguishing COVID-19 from other viral pneumonias that have real time polymerase chain reaction confirmation of the viral agent in a future study.

Furthermore, there is a large commonality in how the lung responds to various pathologies and there is a considerable amount of similarity in the presentation of many diseases in the lung that depend on a plethora of factors (e.g. age, drug reactivity, immune status, underlying co-morbidities). It is likely impossible to differentiate all lung diseases by way of imaging and AI alone, and as such, drastic improvements in positive predictive value are likely difficult to achieve. A multi-omics approach for this is most likely optimal.

Future work is planned to collect additional chest CTs from multiple centres to externally validate the performance of our algorithm. Ultimately, this study focuses on diagnosis whereas prognosis on the future disease trajectory is an even more urgent unmet clinical need that would enable improved resource management (including management decisions regarding the allocation of ventilators). This is the next step for our collaborative research.

## Conclusion

Benchmarked against RT-PCR confirmed cases of COVID-19, our AI framework can accurately detect COVID-19. Thus, providing rapid accurate diagnosis in patients with suspected of COVID-19 infection, facilitating the timely implementation of isolation procedures and early intervention.

## Data Availability

Data is not made available online

## Acknowledgements

The authors acknowledge financial support from Interreg V-A Euregio Meuse-Rhine (Euradiomics project), an ERC advanced grant (ERC-ADG-2015, n° 694812 - Hypoximmuno) and a European Marie Curie grant (PREDICT - ITN - n° 766276) for the execution of this work.

## Online supplement

The lungs were segmented as a single structure using RadiomiX (Oncoradiomics SA, Liège, Belgium) based on convolutional neural networks by combining 3D and 2D architectures. Details on both these architectures are given below.

### 3D lung segmentation

This model architecture consists of a 3D U-Net [1] with residual blocks [2] in the encoder part of the network. Publicly available data from the cancer imaging archive [3] was used to train and validate the model. The specific dataset [4] contains CT scans of 422 confirmed non-small cell lung cancer cases, along with manual segmentations of the left and right lungs. The segmentations were performed by an experienced radiologist and these segmentations were used as a reference standard. The data was randomly partitioned into a training set (n = 322), a tuning set (n = 50), and a test set (n = 50). In order to generate homogeneous CT volumes as input for the model, the following pre-processing steps were performed. All the volumes were resized to 160 × 160 × 448 along the x, y and z axis .and image intensities were clipped at a window width of 1500 HU and a window level of −600 HU (i.e., a standard lung CT window level settings).

The following data augmentations were performed to avoid overfitting [5] on the training dataset:

- Flipping in different directions: up and down, left and right
- Randomly resampling volumes to varying voxel sizes
- Rotating (10–30 degrees) onto the left or right direction
- Reversing the sequence of axial slices

The model was trained with the pre-processed volumes and their corresponding reference labels, using Jaccard loss [6] as an objective function. Here, the loss is calculated in a mini batch of two images per iteration. The network was trained for 10 epochs and at the end of each epoch the Jaccard loss was calculated on the model’s predictions to ensure validation loss convergence.

### 2D lung lobe segmentation

The full lung lobe segmentation model consists of two sub-networks with a similar architecture, but independently trained for the left and right lung (i.e., one model is trained on 2D axial slices for the right upper/middle/lower lobes and the other model is trained on 2D axial slices for left upper/lower lobes). The architecture of the networks is based on a 2D Feature Pyramid Network [7] adapted with ResNext blocks [8] in the encoder. The model was trained and validated on the publicly available subset of the LUNA16 challenge. The subset contains 50 CT volumes manually annotated for lung lobes [9]. Since this is only a small training dataset, the following data augmentation strategies were adopted to increase the training set:

- Resampling to varied voxel sizes and resizing to the common size (160,160,448)
- Cropping and zooming in through left or right lung and resizing the cropped volume to size (160,160,448)
- Zooming out at different ranges
- Rotating left or right at varied degrees

The networks were trained with the 2D axial slices clipped at a window width of 1500 HU and a window level of −600 HU and with their corresponding reference labels. The networks’ weights were updated by using the Adam optimizer at an initial learning rate of 1e-5 [10]. The model was trained using customized focal Tversky loss [11] (adapted for calculating loss over multiple classes) as an objective function where the loss is calculated in a mini batch of four images per iteration. The network was trained for 5 epochs and at the end of each epoch, the customized focal Tversky loss was calculated on the model’s predictions to ensure validation loss convergence.

The predicted segmentations of each architecture (i.e., the segmentation output from both the 3D and the 2D segmentation models) are ensembled and the intersection constitutes the final total lung segmentation which is used for extraction of radiomics features. The deep learning-based lung segmentation achieved a mean Dice similarity coefficient score of 0.86 across the publicly available datasets which indicates adequate precision (i.e. no significant over or under segmentation).

